# The quarantine hospital strategy as a way to reduce both community and nosocomial transmission in the context of a COVID-like epidemic

**DOI:** 10.1101/2025.01.13.25320454

**Authors:** Théo Pinettes, Quentin J. Leclerc, Kévin Jean, Laura Temime

## Abstract

Nosocomial infections of both patients and healthcare workers (HCWs) in hospitals may play an important part in the overall dynamics of a viral pandemic, as evidenced by the recent COVID-19 experience. A strategy to control this risk consists in dedicating some hospitals to the care of infected patients only, with HCWs alternating between shifts of continuous stay within these hospitals and periods of isolation. This strategy has been implemented locally in various settings and generalized in Egypt. Here, using a mathematical model coupling hospitals and community, we assess the impact of this strategy on overall epidemic dynamics. We find that quarantine hospitals may significantly reduce the number of cumulative cases, as well as the peak incidence. These benefits are highest when effective control strategies are in place in the community and symptomatic HCWs comply with self-isolation recommendations. Our results, which are robust to variations in assumed biological characteristics of the virus, suggest that the quarantine hospital strategy should be considered in future pandemic contexts to best protect the entire population.

## Introduction

When facing a pandemic transmissible agent, isolating infected individuals is generally one of the first measures implemented. If disease symptoms often require hospitalization, hospitals may become clusters of infectious individuals and play an important role in overall epidemic dynamics, through both patient-to-patient and patient-to-healthcare workers (HCWs) transmission [1, 2]. Hence, restraining contacts between hospitals and the rest of the community may help protect the entire population. In the context of the COVID-19 pandemic, diverse initiatives based on this observation emerged as early as spring 2020, such as some French nursing homes isolating their staff with the residents for several weeks [3]. However, the most elaborate strategy was the Egyptian one. Under WHO supervision, the country redesigned its entire healthcare model to control risks of viral spread from hospitals to community. The government assigned certain hospitals to care only for externally-referred COVID-19 patients, making other hospitals of the same area as free of the disease as possible. These so-called “quarantine hospitals” had rotating medical teams residing continuously in the hospital during 1 or 2 week-long working shifts [4]. Here, we consider this unique healthcare model as an approach that may be generalized to other countries and pandemic contexts – provided that the circulating pathogens are to some extent biologically similar to SARS-CoV-2 – and refer to it as the “quarantine hospital strategy”.

Recent studies have assessed the epidemiological [4] and psychological [5] consequences incurred by health care workers (HCWs) working in quarantine hospitals. Briefly, results suggest that whether COVID-19 acquisition or mental health risk for quarantine hospital HCWs exceeds those of classical settings depends on the amount of resources deployed to set up these quarantine hospitals. However, while the benefits of the quarantine hospital strategy in terms of epidemic containment are widely assumed, they have never been quantified, nor have they been weighed against the risks they may also carry, such as a possible increased exposure for HCWs.

Significant organizational differences have been observed across Egyptian quarantine hospitals [4], suggesting that the exact protocol to maximise the benefits of the quarantine hospital strategy is not yet known. More generally, in anticipation of an upcoming COVID-19 wave or even a new pandemic, it would be valuable to provide insight into the range of conditions where the quarantine hospital strategy could be an efficient response, including biological characteristics of the virus, effectiveness of implemented control measures in the community and standard hospitals, shift duration within quarantine hospitals, etc.

To address these questions, we developed a novel stochastic compartmental model to simulate the propagation of a SARS-CoV-2-like virus among the population of a medium-sized city hosting one quarantine hospital and one other hospital referred to as “usual” hospital (Fig 1). The model accounts for patients and HCWs circulating between the community and the different hospitals, as well as for different transmission routes across populations (HCWs and non-HCWs) and settings (community, usual and quarantine hospitals). By comparing outputs from model simulations performed with a quarantine hospital (“quarantine hospital strategy”) and without a quarantine hospital (“reference strategy”), we assessed potential gains of the quarantine hospital strategy in terms of burden and infection peak reduction during a typical COVID-19-like outbreak. We also evaluated the impact of the strategy on HCWs specifically. We further explored the resilience of the strategy to more diverse contexts by performing widerange sensitivity analyses.

**Figure 1:**
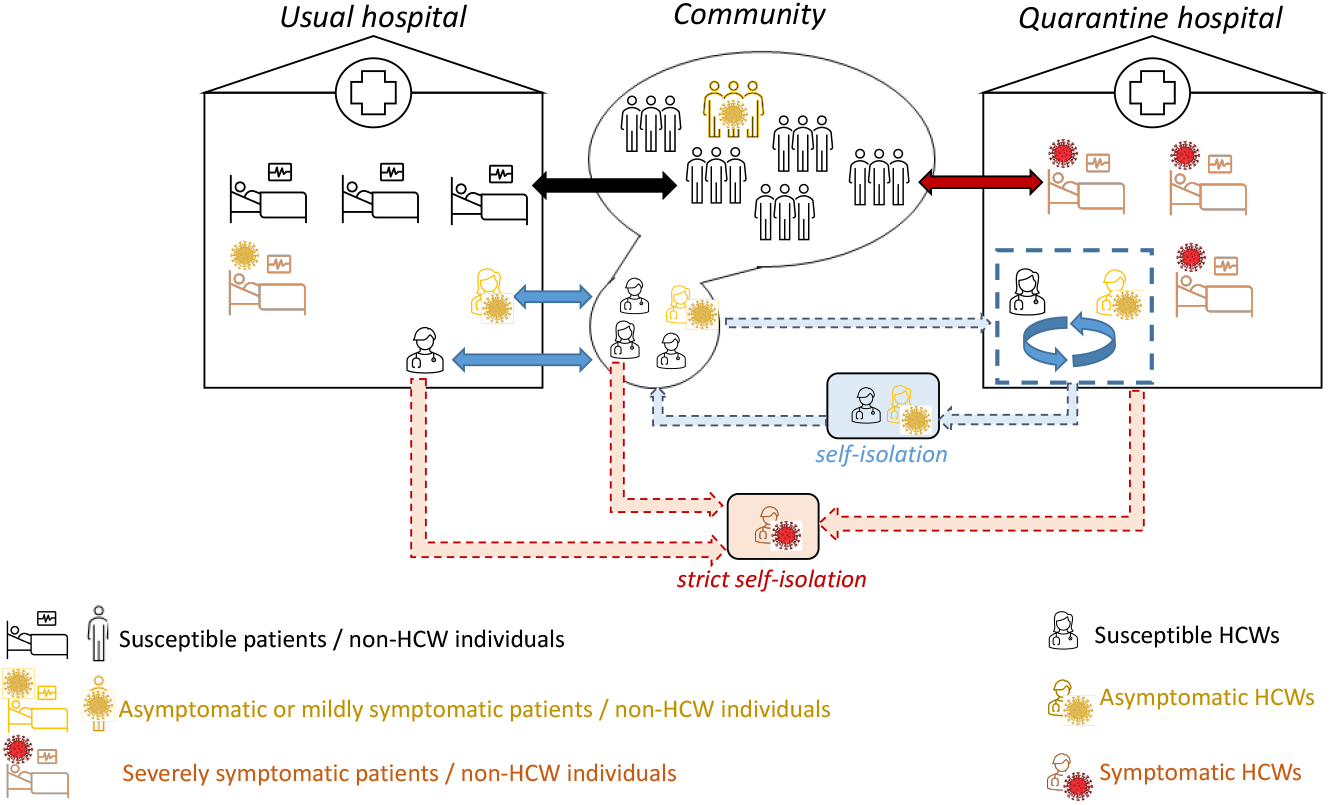
Global model structure: movements of individuals (non-HCWs and HCWs) between the community, quarantine and usual hospitals.

## Results

### The quarantine hospital strategy may significantly reduce the overall epidemic burden

In our main analysis, we assumed a community transmission rate of the virus of 0.107 days^−1^, corresponding to a basic reproduction number in the community, *R*_*C*_, of 1.05. This reproduces the scenario of an epidemic wave with control measures in place in the community, a relevant context for implementing the quarantine hospital strategy. We also calibrated the within-quarantine hospital transmission rate to HCWs at 0.112 days^−1^, allowing to reproduce the highest acquisition risk measured in Egyptian quarantine hospitals, at 48% per week, in an earlier study [4]. All other parameter values for the baseline scenario we present here are provided in Materials and Methods (see Table 2). In this scenario, the quarantine hospital strategy flattens the epidemic curve. Amongst non-HCWs, the infection prevalence peak is delayed by more than 50 days and decreased from 3.7% (2.5% - 4.3%) to 1.9% (1.1% - 2.5%) (Fig 2a).

**Figure 2:**
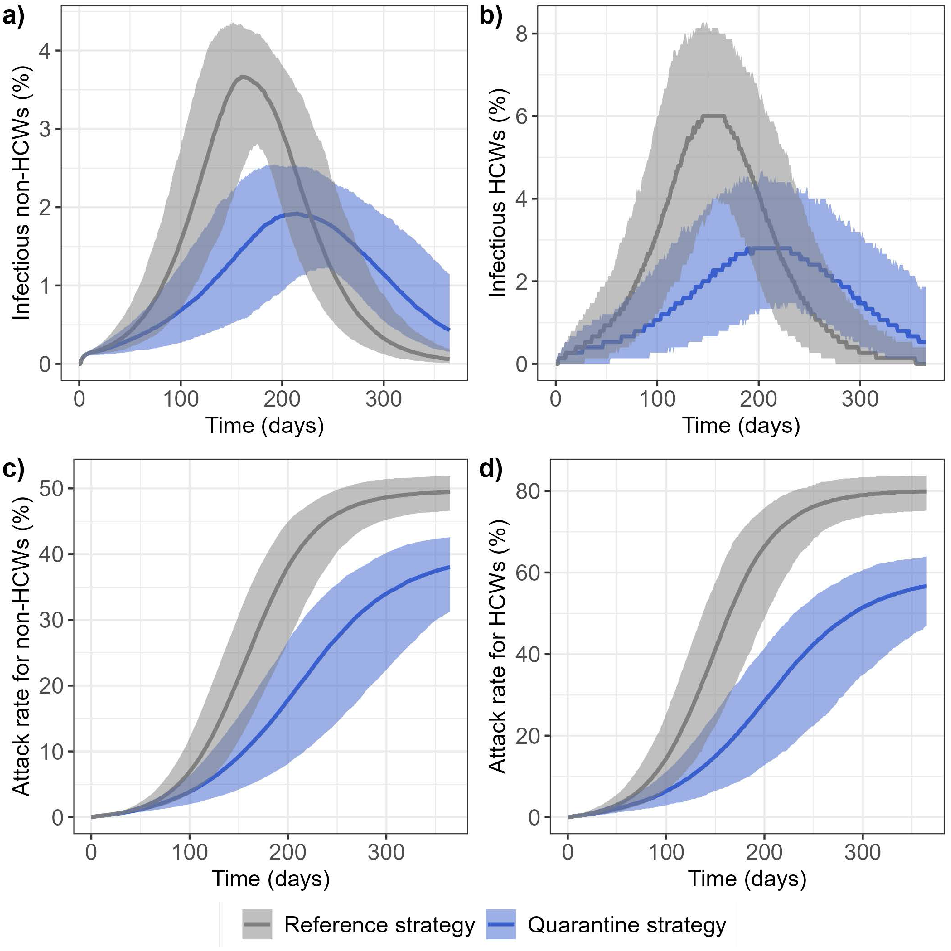
Epidemic curves comparing the reference (grey) and quarantine (blue) strategies. Predicted changes over 365 days in the median and 95% prediction band of the percentage of infectious individuals (I and A compartments) amongst a) non-healthcare workers and b) healthcare workers. Attack rate in c) non-healthcare workers and d) healthcare workers.

As a result, the overall epidemic burden is reduced, with a decrease in the attack rate after 365 days of 11.4 (9.4 - 15.3) percentage points amongst non-HCWs (Fig 2c).

### HCWs play a key role in virus spread in the community

In Figure 3 we present, with or without quarantine hospitals, the proportion of total number of new infections in the community acquired from non-HCWs, from HCWs resting between work shifts, from symptomatic isolated HCWs or from HCWs isolated after a shift in a quarantine hospital. Our predictions suggest that HCWs are an important vector of virus spread in the community, especially symptomatic isolated HCWs, who, even when assumed to have a 80% reduced risk of onward transmission due to their isolation, may be at the source of almost 25% of all acquisition events in the community over the 365 days.

**Figure 3:**
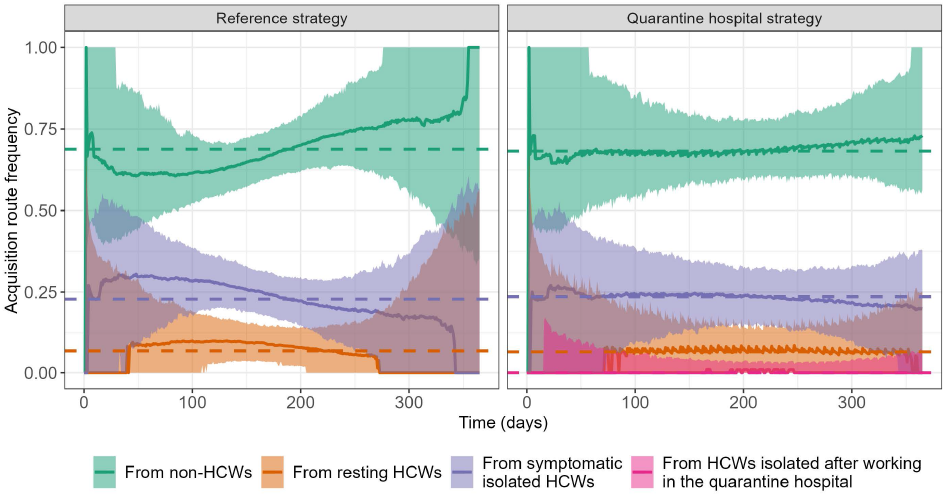
Distribution of acquisition routes over time for non-HCW infections in the community, comparing the reference (left) and the quarantine hospital (right) strategies. Time changes in the median proportion of new infections is depicted over 365 days (solid lines), along with the 95% prediction band (shaded areas), for each route: acquisitions from non-HCWs, from HCWs resting between work shifts, from symptomatic HCWs strictly isolating in the community and, in the quarantine hospital strategy case, from HCWs isolating after a shift in the quarantine hospital. The dashed lines show the median acquisition frequency over the entire 365 days.

The overall distribution of acquisition sources does not substantially differ between the reference and quarantine hospital strategy (Fig 3, dashed lines). HCWs with no symptoms who self-isolate after their working shift in the quarantine hospital play a minor role in the epidemic propagation, as their median contribution to the daily community infections throughout the whole wave does not exceed 1%.

### The quarantine hospital strategy dramatically reduces nosocomial acquisitions in hospitalized patients

Unsurprisingly, model-predicted nosocomial COVID-19 acquisitions collapse thanks to the quarantine hospital strategy, with an 89.8% (85.8% - 92.2%) reduction in nosocomial acquisition frequency (Table 1).

**Table 1:**
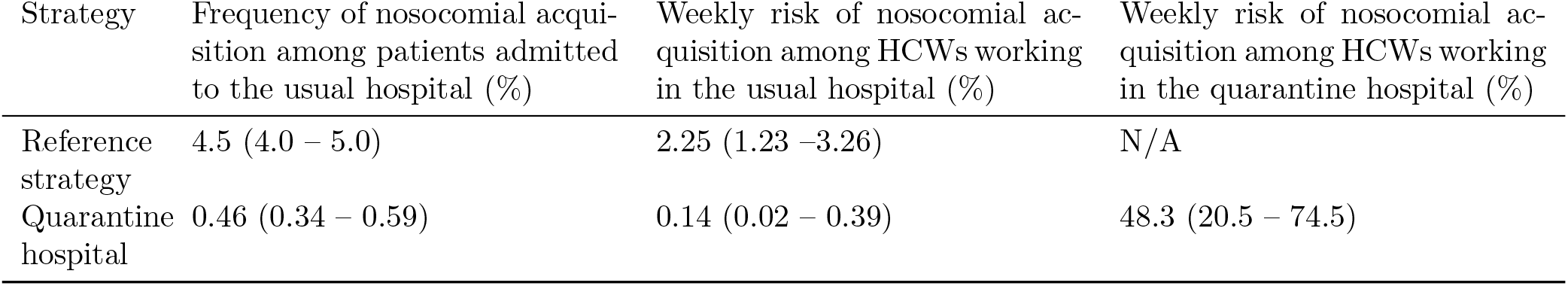
Comparison of the frequency of nosocomial acquisitions among patients and HCWs in the two strategies. The table provides, for patients, the average risk of nosocomial acquisition over their entire hospital stay; and for HCWs, the average weekly risk of occupational acquisition.

**Table 2:** Model parameters.

| Symbol | Denomination | Value | Range explored | Source |
| --- | --- | --- | --- | --- |
| $T_E$ | Mean incubation duration | 4.6 days | $\mathcal{N}(5, 2) \cap [0, \infty[$ | [20] |
| $T_I$ | Mean infectious duration | 9.8 days | $\mathcal{N}(9.5, 3) \cap [0, \infty[$ | [21] |
| $\varphi$ | Proportion of asymptomatic cases | 20% | $\mathcal{N}(0.2, 0.02) \cap [0, 1]$ | [18] |
| $\psi$ | Proportion of severe cases (who may require hospitalization) | 7% | $\mathcal{N}(0.07, 0.02) \cap [0, 1]$ | [12] |
| $\kappa$ | Infectiousness reduction in asymptomatics | 32% | $\mathcal{N}(0.32, 0.07) \cap [0, 1]$ | [18] |
| $\beta_C$ | Transmission rate in the community | 0.107 days<sup>-1</sup> | $\mathcal{N}(0.107, 0.0107) \cap [0, \infty[$ | - |
| $\beta_{1ref}$ | Transmission rate in usual hospitals in the reference strategy | 0.117 days<sup>-1</sup> | $\mathcal{N}(0.117, 0.0117) \cap [0, \infty[$ | [22] |
| $\beta_{1Q}$ | Transmission rate in usual hospitals in the quarantine hospital strategy | 0.112 days<sup>-1</sup> | $\mathcal{N}(0.112, 0.0112) \cap [0, \infty[$ | [22] |
| $\beta_2$ | Transmission rate in quarantine hospitals | 0.122 days<sup>-1</sup> | $\mathcal{N}(0.122, 0.0122) \cap [0, \infty[$ | [4] |
| $N_{tot}$ | City size | 50000 | - | - |
| $N_{hcw}$ | Number of healthcare workers in the city | 750 | - | - |
| $R_{ph}$ | Patients-to-HCW ratio in all hospitals | 3 | - | - |
| $\epsilon$ | Symptomatic HCW degree of non-compliance with self-isolation | 20% | $\mathcal{U}[0, 40\%]$ | - |
| $\epsilon_{H2}$ | Asymptomatic quarantine hospital HCW degree of non-compliance with self-isolation | 50% | $\mathcal{U}[40\%, 70\%]$ | - |
| $T_1$ | Average patient length of stay in the usual hospitals | 5.4 days | - | [23] |
| $T_Q$ | Average patient length of stay in the quarantine hospitals | 9.8 days | - | - |
| $T_2^{shift}$ | Working shift duration in quarantine hospitals | 7 days | $\mathcal{U}[7;14]$ | [4] |

### On average, HCWs are not put at excess risk by the quarantine hospital strategy

The quarantine hospital strategy delays the infection prevalence peak in HCWs by more than 50 days and decreases it from 6.0% (3.9% - 8.1%) to 2.8% (1.3% - 4.4%) of the total population (Fig 2b). As a result, the specific epidemic burden of HCWs is reduced, with a decrease in the attack rate after 365 days of 23.2 (19.7 - 28.3) percentage points (Fig 2d) in the quarantine hospital strategy. This reflects the fact that benefits for HCWs working in usual hospitals or in the community more than compensate the increased risk to HCWs working in quarantine hospitals (Table 1). Notably, the risk of occupational contamination is reduced by 94% in usual hospitals, even though our model predicts a 48% weekly risk of acquisition for HCWs within quarantine hospitals, a worst-case hypothesis based on observations in Egyptian hospitals [4].

### The quarantine hospital strategy is most effective when epidemic control is high in the community and symptomatic HCWs comply well with self-isolation

Figure 4 depicts the predicted reduction, compared to the reference strategy, in the total epidemic burden over 365 days (Fig 4a) and in the incidence at the epidemic peak (Fig 4b) as a function of *R*_*C*_, the basic reproduction number in the community, and *ϵ*, the degree of non-compliance to self-isolation in symptomatic HCWs. Obtaining a reduction from the quarantine hospital strategy larger than 50% requires both stringent control measures in the community (e.g., a general population lockdown) and for symptomatic HCWs to comply with self-isolation (*R*_*C*_ ≤ 1 and *ϵ* ≤ 0.1). Such conditions also lead to a substantial infection peak reduction (approx. 60%), but the quarantine hospital strategy provides important improvements in that respect even in a context of less strict control measures (e.g., 45% peak size reduction when *R*_*C*_ ≤1.1 and *ϵ* ≤0.15). In addition, the specific epidemic burden and infection peak for HCWs are always lower in the quarantine hospital strategy than in the reference strategy, irrespective of assumed values for *R*_*C*_ and *ϵ* (Fig 4, right panels).

**Figure 4:**
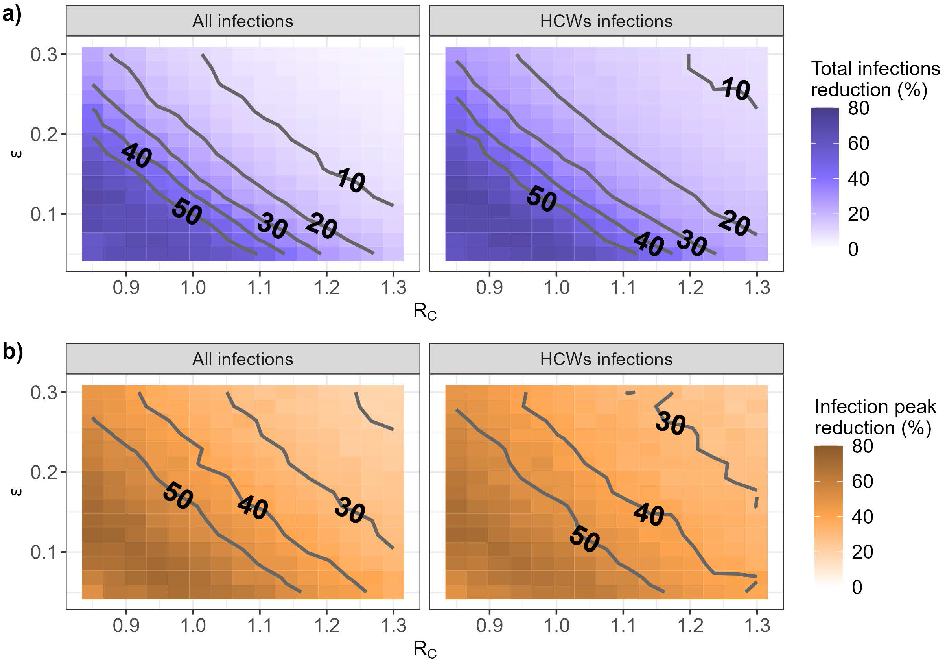
Predicted reduction achieved by the quarantine hospital strategy (as compared with the reference strategy) in (a) the cumulative number of infections over 365 days and (b) the peak incidence, as a function of *R*_*C*_, the basic reproduction number in the community, and *ϵ*, the level of self-isolation non-compliance in symptomatic HCWs.

Finally, irrespective of *R*_*C*_ and *ϵ*, the reduction in nosocomial acquisitions remains above 80% (Fig 5).

**Figure 5:**
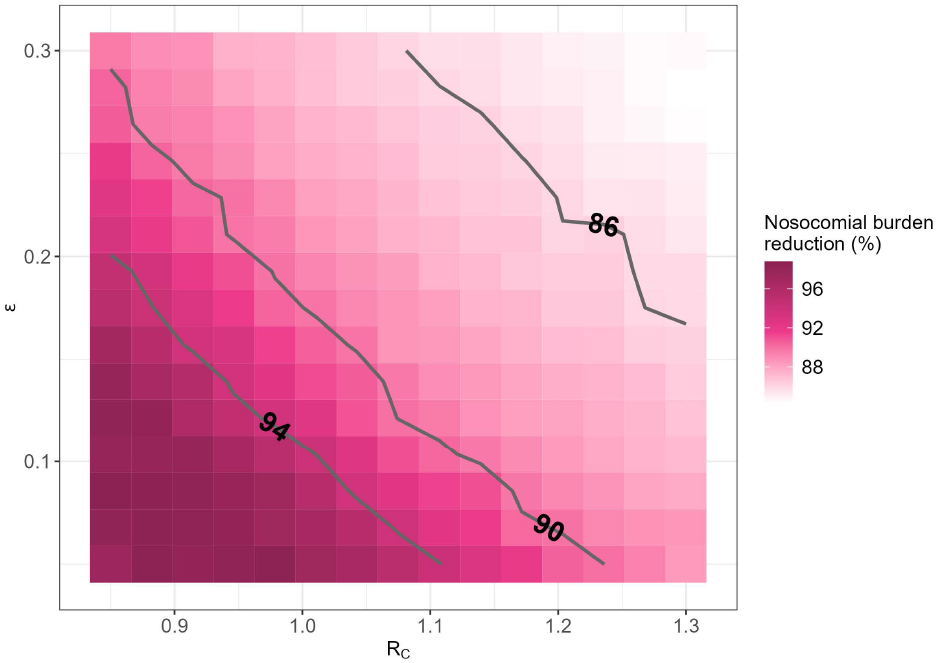
Reduction in the cumulative number of nosocomial acquisitions over 365 days, as a function of assumed values for *R*_*C*_, the basic reproduction number in the community, and *ϵ*, the level of self-isolation non-compliance in symptomatic HCWs.

### Benefits of the quarantine hospital strategy are robust to parameter variability

By conducting a partial rank correlation analysis (Fig 6), we identified that the two parameters which most impact epidemic dynamics were *β*_*C*_ and *ϵ*, the two key factors explored above. All hospital policy-related parameters other than *ϵ* play a minor role in epidemic containment.

**Figure 6:**
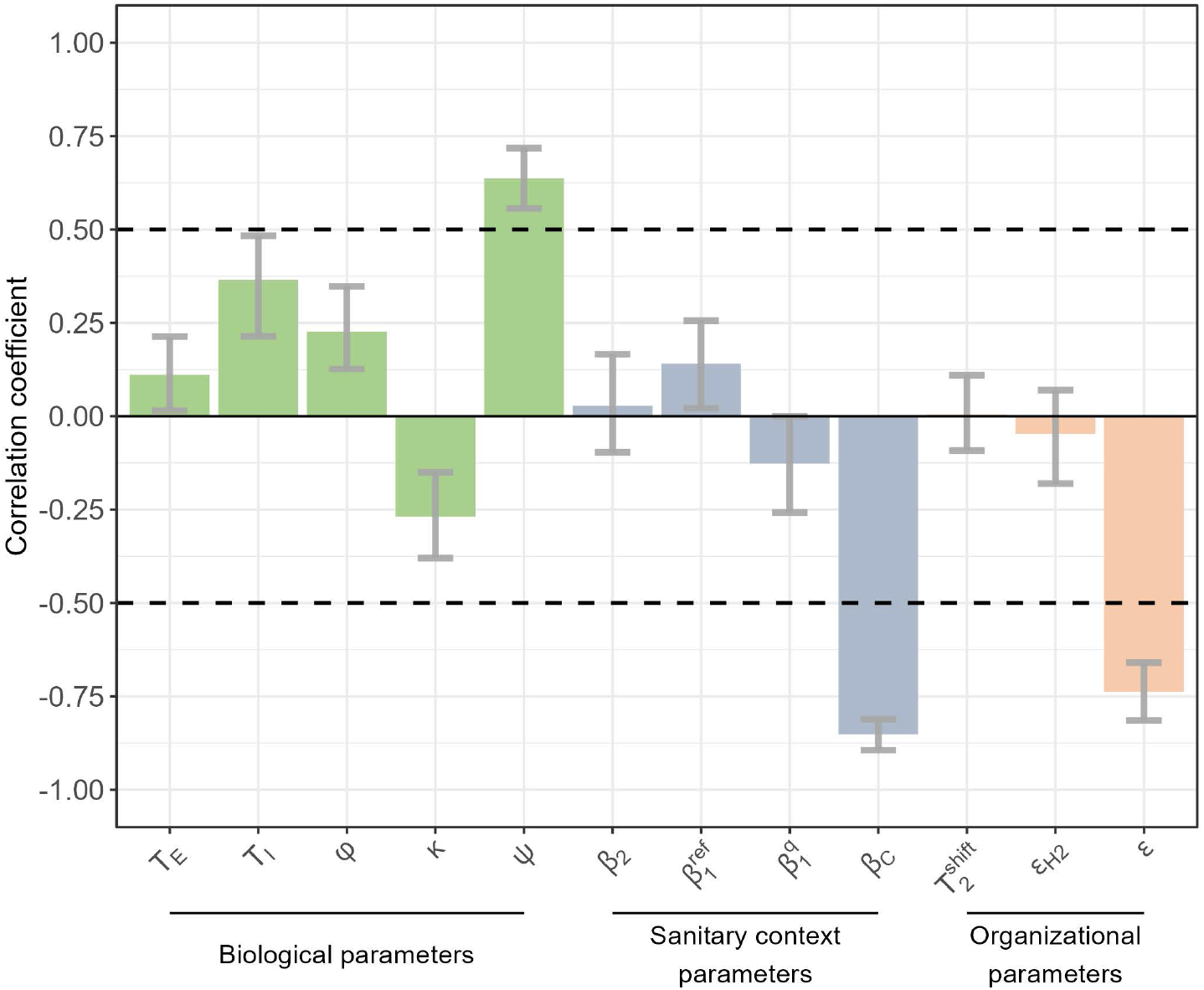
Sensitivity analysis: partial rank correlation coefficients (PRCC) between the predicted reduction in the cumulative number of cases over 365 days and model parameters. Three categories of parameters are explored: biological characteristics of the virus, parameters resulting from the population structure and implemented control measures within the usual hospitals and community (“sanitary context parameters”) and parameters describing the implementation of the quarantine hospital strategy (“organizational parameters”). The full list of model parameters is provided in Table 2, along with their baseline values and explored ranges.

Biological variability may also affect our results, with notably higher predicted effectiveness of the quarantine hospital strategy when the proportion of severe cases *ψ* increases.

Finally, assumed values for within-hospital transmission rates 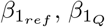 and *β*_2_ have a low impact on our predictions (Fig 6).

## Discussion

The modelling carried out in this study provides evidence on the conditions under which the quarantine hospital strategy can help contain epidemics. For a virus similar to SARS-CoV-2, the two key conditions identified are: (i) some control measures must be in place in the community (expressed by the *R*_*C*_ parameter); and (ii) symptomatic HCWs must rigorously comply with self-isolation (expressed by the *ϵ* parameter). In conditions similar to those observed during the second COVID-19 epidemic wave, the quarantine hospital strategy induces a 43.3%(24.6 – 59.9) infection peak reduction and a 22.0%(12.6 – 35.1) virus burden reduction; additionally, it reduces nosocomial infections by more than 89.8% (85.8% - 92.2%), thus substantially reducing the risk among the most vulnerable individuals. The strategy efficacy may vary slightly with the transmission context in hospitals, but significant epidemic containment benefits hold as long as the key conditions are respected.

### Validation

Our identification of the basic reproduction number in the community as a key determinant of success for the quarantine hospital strategy is consistent with the conclusions of a recent British study [2], which underlined the major contribution of nosocomial transmission in the overall COVID-19 burden during lock-down periods. This is notably because measures implemented in the community, such as a lockdown, barely reduce transmission inside hospitals – medical care requires close contacts regardless of distancing measures adopted in the community, thus the relative contribution of hospitals to epidemic spreading increases during periods of lockdown. Hence, in a context of strong control measures, mitigating virus propagation within hospitals is particularly relevant, and that is precisely the goal of the quarantine hospital strategy. Our analysis highlights HCWs as important vectors of transmission among the community, hence why we found the compliance of HCWs to self-isolation to be critical for the success of the quarantine hospital strategy.

Unfortunately, while data from Egyptian quarantine hospitals has allowed us to quantify the risk for HCWs [4], no data is available to quantify SARS-CoV-2 spread between hospitals and the general population in Egypt, therefore preventing the full calibration of our model and/or validation of its predictions. However, in the absence of observational data, modeling is the only way to gain insight on the potential benefits of the quarantine hospital strategy. In addition, our model predicts an acquisition risk of 48% per week for HCWs assigned to quarantine hospitals that lies within the range previously estimated from data observed in Egyptian quarantine hospitals (from 13% per 14-day shift to 48% per 7-day shift) [4]. We chose to calibrate the transmission rate within quarantine hospitals to reproduce a risk at the upper bound of this range, in order to be as conservative as possible when assessing potential benefits of the quarantine hospital strategy. Finally, our sensitivity analysis highlighted that the assumed value for this within-hospital transmission rate has a limited impact on the predicted benefits of the quarantine hospital strategy at the entire population level.

### Limitations

The sensitivity analysis we performed underlined the impact on our results of the assumed transmission rate at the community level *β*_*C*_, as well as, to a lesser extent, of the assumed proportion of severe cases. This could have implications in case of emergence of a new variant with significantly different transmission potential, or when accounting for vaccination. However, as evidenced by the analyses presented in Figures 4 and 5, our main conclusions hold, provided that *R*_*C*_ is not too high - a condition which should only be reinforced in the presence of widespread vaccination.

Our model does not account for age-specific patterns in terms of susceptibility, mixing and eventually contribution to disease transmission. Previous studies documented the role of age in the probability of asymptomatic infection or hospitalization [6, 7, 8]. Similarly, contact patterns are documented to be age-dependent [9, 10]. Therefore, the quarantine hospital strategy is expected to result in the isolation of older patients, more likely to show symptoms and with higher level of infectiousness, but with lower contact rates in average. Results obtained in other settings suggested that the contribution of each age group to transmission was highest among the 20-29y old, and then decreased with age [11]. If such a pattern was confirmed, disregarding age may have led us to overestimate the quarantine hospital strategy impact, because isolation may in reality disproportionately affect age groups that contribute slightly less to transmission.

Additionally, our model does not account for HCW-to-HCW transmission. This may significantly increase the risk within hospitals, especially in quarantine settings where HCWs may sometimes share resting and conviviality rooms. Hence, proposing infectious control recommendations specifically targeted at interactions between HCWs may be key in quarantine hospitals.

We also neglected hospitalization of HCWs with severe symptoms. Since this would only concern around 7% of infected HCWs (*ψ* parameter [12]), while HCWs themselves only represent 1.5% of the population considered, this should not have a major impact on our results.

Finally, we did not include diagnostic tests in our analysis, and instead reproduced the situation observed in Egypt before reliable rapid tests became available [4, 13, 14]. We assumed that all severely symptomatic individuals are correctly identified as COVID-19 cases and hospitalised in the quarantine hospital. Although we believe this is reasonable since these cases are less likely to be undetected, we acknowledge that in reality some severe cases could be treated as false negatives and remain in the usual hospital, thereby decreasing the effectiveness of the quarantine hospital strategy. Conversely, we assumed that none of the asymptomatic or mildly symptomatic individuals are hospitalized for COVID-19. If these individuals could instead be identified, we expect this would reduce transmission in the community and therefore further increase the benefits of the quarantine hospital strategy, since the relative importance of nosocomial and HCW-mediated transmission would increase. Since we did not consider tests, we also assumed that all HCWs self-isolated after their shift in the quarantine hospital, with a compliance level of *ϵ*_*H*2_ regardless of whether they were uninfected or infected asymptomatic. Reliable exit testing after a shift would instead better distinguish between truly infected HCWs who need to isolate and uninfected HCWs who do not, thereby reducing the risk of HCW-mediated transmission in the community and improving the effectiveness of the quarantine hospital strategy.

### Public health implications

Our results suggest that the quarantine hospital strategy has potential major public health benefits for both the control of SARS-CoV-2 and a range of other biologically similar viruses, as evidenced by the sensitivity analysis we performed. In particular, beyond the overall reduction in cases we predicted, two key additional impacts may be underlined.

First, nosocomial acquisitions would be dramatically reduced by the quarantine hospital strategy. As hospital patients tend to be older and more fragile than the general population [15], this means that the mortality benefits of the quarantine hospital strategy would be larger than its morbidity benefits.

Second, the quarantine hospital strategy tends to flatten the epidemic curve (Fig 2a), thus allowing for better preparedness and decreased risk of saturation of the healthcare system, a key issue in many countries worldwide during the COVID-19 pandemic [16].

### Feasibility

In addition to demonstrating the potential benefits of the quarantine hospital strategy, the present study provides some insight to guide its implementation. First, despite exposing some HCWs to highly contagious residents, the quarantine hospital strategy reduces the risk of COVID-19 contamination for HCWs overall, suggesting that benefits to HCWs in usual hospitals and the community more than compensate the increased risk to HCWs in quarantine hospitals. As the transmission rate we assumed within quarantine hospitals was actually in the higher range of previous estimates from Egypt [4], the actual overall risk for HCWs may be even lower than our prediction. Second, while one may expect a negative impact of working in a quarantine hospital on HCW mental health and well-being, a study performed in different Egyptian hospitals actually found a lower risk of depression in quarantine-hospital HCWs, as compared with HCWs working in standard frontline hospitals [17]. In addition, the sensitivity analysis we performed shows that quarantine working shift duration as well as the respect of self-quarantine after working shifts are not crucial for the strategy efficiency. Hence, one-week-long working shifts, which are almost as effective as two-week-long shifts, could be favored, and “relaxed” self-isolation after these shifts (e.g. contact limitation to close relatives) could be recommended in asymptomatic HCWs.

## Conclusion

In conclusion, we have described the potential benefits of the quarantine hospital strategy and identified the optimal conditions to maximize these benefits during an epidemic situation. From our findings, we propose a series of recommendations for implementing such a strategy when facing a virus with characteristics similar to those of SARS-CoV-2, namely a significant fraction of cases requiring hospitalization, incubation and infectiousness durations of approximately one week, a limited proportion of asymptomatics and a reduction of infectiousness in asymptomatics.

First, quarantine hospital strategies must be paired with control measures to limit viral propagation in the community (e.g., non-pharmaceutical interventions or vaccination), translating as an effective reproduction number below 1.1. Second, symptomatic HCWs need to engage in strict self-isolation, with the aim of a more than 90% reduction in their contacts. Third, when organizing quarantine hospitals, one-week-long working shifts for HCWs, followed by a self-isolation of the same duration, should be preferred.

These recommendations go beyond what was implemented in the Egyptian example. They pave the way for complementary work on the subject, such as evaluating the mental health, logistic burden and economical cost of such a healthcare reorganisation. This would require answering a number of yet unaddressed questions, including for instance the maximal duration that HCWs are ready to respect the constraints associated with quarantine hospitals, the criteria to designate the settings that will become quarantine hospitals, the costs associated with equipment transfers between usual hospitals and quarantine hospitals – e.g., respiratory aid equipment. Collecting data on these questions and performing a cost-effectiveness analysis would be useful for the quarantine hospital strategy to become a valid and well-documented option for health-policy makers in epidemic situations.

## Methods

### The quarantine hospital strategy

We developed a compartmental stochastic model to simulate the propagation of a SARS-CoV-2-like infectious agent among the population of a medium-sized city hosting a single quarantine hospital and a single “usual” hospital. HCWs and other individuals (potential patients) circulate between the community and the two hospitals (fig 1).

In each subpopulation and setting, individuals are distributed into 5 different compartments depending on their infection status: Susceptible, Exposed, Infectious symptomatic, Infectious asymptomatic or Recovered.

Susceptible (S) individuals may become infected by an infectious virus carrier and enter the incubation stage (E), during which they are not infectious yet. Individuals exit the incubation stage at rate 1*/T*_*E*_, and either become Infectious symptomatic or Infectious asymptomatic. For non-HCWs, mildly symptomatics are grouped together with asymptomatics in the Infectious asymptomatic compartment (*I*_*M*_), which is reached with probability 1 − *ψ*. This leaves only severely symptomatic non-HCWs, who will systematically be hospitalized, in the Infectious symptomatic compartment (*I*_*H*_), which is reached with probability *ψ*. Conversely, for HCWs, all symptomatically infectious individuals are included in the Infectious symptomatic compartment (*I*_*S*_), which is reached with probability 1 − *φ*, and the Infectious asymptomatic compartment (*I*_*A*_), which is reached with probability *φ*, only includes true asymptomatic HCWs. Both symptomatics and asymptomatics recover at rate 1*/T*_*I*_, entering the Recovered (R) compartment.

The force of infection (probability of the S-to-E transition, detailed in the SI appendix) depends on several factors:

- The time-dependent proportion of infectious individuals in the population; noting that infectiousness is reduced by a factor *κ* in asymptomatic individuals [18];
- The transmission rate *β*, that accounts for both the rate of contacts within and between the different sub-populations (which is reduced by mandatory self-isolation), and the per-contact probability of infection between susceptible and infectious individuals (which is reduced by contact precautions implemented, e.g., masks). Three distinct transmission rates were defined depending on the setting: *β*_*C*_ within the community, *β*_1_ within usual hospitals, and *β*_2_ within quarantine hospitals.

Non-COVID-19 hospitalized patients leave the usual hospital at rate 1*/T*_*h*_, and admission of patients coming from the community is adjusted so that the number of hospitalized patients remains within total hospital capacity (fig S1 in the SI appendix). All individuals except HCWs, including patients hospitalized in the usual hospital, are assumed to be admitted into the quarantine hospital as soon as they become severely symptomatic (*I*_*H*_ compartment) and to remain there until they recover, whereupon they return to the community. In other words, we assume that the hospital length of stay for patients with COVID-19 is equal to their infectious duration of 9.8 days (Table 2), which falls within the range of estimated length of stay in Africa [19].

HCWs working in the usual hospital follow daily rotations: one third of the HCWs leave the community each morning, joining the usual hospitals, and return to the community on the evening. Regarding the quarantine hospital, rotations occur on a 1-week basis, depending on the quarantine shift duration 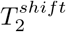 ; enough HCWs to respect the patients-to-HCW ratio *R*_*ph*_ in the quarantine hospital enter the hospital from the community after each time period and remain there for this duration 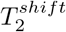 [4]. At the end of their working shift, they leave the quarantine hospital and self-isolate in the community for an additional period of equal duration [4]. During self-isolation, their contribution to the force of infection is reduced by a factor (1 −*ϵ*_*H*2_)∈ (0, 1). After that period, they return to the community.

In addition, HCWs who become infectious and symptomatic are assumed to strictly isolate in the community, irrespective of their original location. During this strict isolation, their contribution to the force of infection is reduced by a factor (1− *ϵ*). HCWs in strict isolation return to the community when they recover.

All model parameters are listed in Table 2, along with their definition and assumed baseline values. Baseline values were chosen to reproduce a typical European city facing COVID-19 between September 2020 and end of December 2020. As an epidemic wave with control measures, this period is representative of a relevant context for implementing the quarantine hospital strategy. Parameter distributions were chosen to account for variability around these baselines values wide enough to include the parameters of a COVID-19 variant or a new virus, various disease propagation scenarios among the community and in hospitals, and to represent different plausible healthcare organization choices.

Formulas for the forces of infection are provided in the SI appendix, along with a figure depicting the successive possible states for HCWs and non-HCWs. The model was implemented using R version 4.2.1 and the odin package (https://cran.r-project.org/web/packages/odin/odin.pdf). The code is available in a dedicated GitHub repository (https://github.com/MESuRS-Lab/quarantine_strategy).

### The reference strategy

The quarantine hospital strategy is compared to a reference strategy in which there is no quarantine hospital. This is modelled as above for the community and usual hospital. Symptomatic individuals are admitted into the usual hospital, except for HCWs who are still assumed to strictly self-isolate, in which case the level (1 − *ϵ*) of reduction in their contribution to the force of infection is the same as in the quarantine hospital strategy. In this strategy, the usual hospital hosts all hospitalized infected individuals in addition to other patients. As the total hospital capacity remains constant to preserve the overall patient-per-HCW ratio, this implies that over time, in the quarantine hospital strategy, less individuals are hospitalized in the usual hospital to compensate for individuals hospitalized in the quarantine hospital (fig S1 in the SI appendix), reflecting the deprogramming of non-urgent hospitalizations that was observed during the COVID-19 pandemic. In addition, to reflect the disorganization induced by infected patients, the transmission rate within the usual hospital is increased to 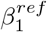.

### Scenario assessment

To assess epidemic containment allowed by the quarantine hospital strategy for a given set of parameters, the present study compares the median epidemic curves obtained over 1,000 simulations for each scenario, focusing on the following indicators : i) the cumulative number of infections over 365 days and ii)the maximum daily number of new infections (incidence peak). Both were assessed in the community and in the HCW population.

### Sensitivity analysis

Lastly, in order to identify the most influential parameters, we performed a sensitivity analysis. The parameter space was explored through Latin Hyper-cube Sampling (LHS) and Partial Rank Correlation Coefficients (PRCC) between each parameter and the relative reduction in the cumulative number of infections over 300 days brought by the quarantine hospital strategy. This analysis was performed using the LHS R-package with 150 parameter samples based on the parameters prior laws. PRCC confidence intervals were computed from 100 bootstrap samples.

## Data Availability

The code is available in a dedicated GitHub repository (https://github.com/MESuRS-Lab/quarantine_strategy)

