## Supplementary Information for "The quarantine hospital strategy as a way to reduce both community and nosocomial transmission in the context of a COVID-like epidemic"

January 13, 2025

#### Model diagram

Figure 1 depicts possible infection statuses for HCWs and non-HCWs: susceptibles (S), exposed (E), infectious (I) and recovered (R). Infectious individuals are further stratified depending on their symptoms, with asymptomatic and mildly symptomatic individuals grouped together in non-HCWs, while all symptomatic HCWs are grouped together.

#### Transmission rates

Transmission rates $\beta_X^{ij}$ in a setting X (denoted as C for the community, 1 for the usual hospital and 2 for the quarantine hospital) from individuals of category i to individuals of category j (category denoted as h for healthcare workers and p for non-healthcare workers) are computed as:

##### In the commmunity

$$\beta_C^{pp} = \beta_C \tag{1}$$

$$\beta_C^{ph} = \beta_C \tag{2}$$

$$\beta_C^{hh} = \beta_C \tag{3}$$

$$\beta_C^{hp} = \beta_C \tag{4}$$

The equivalent basic reproduction ratio $R_C$ is computed as $R_C = \beta_C \times T_I$

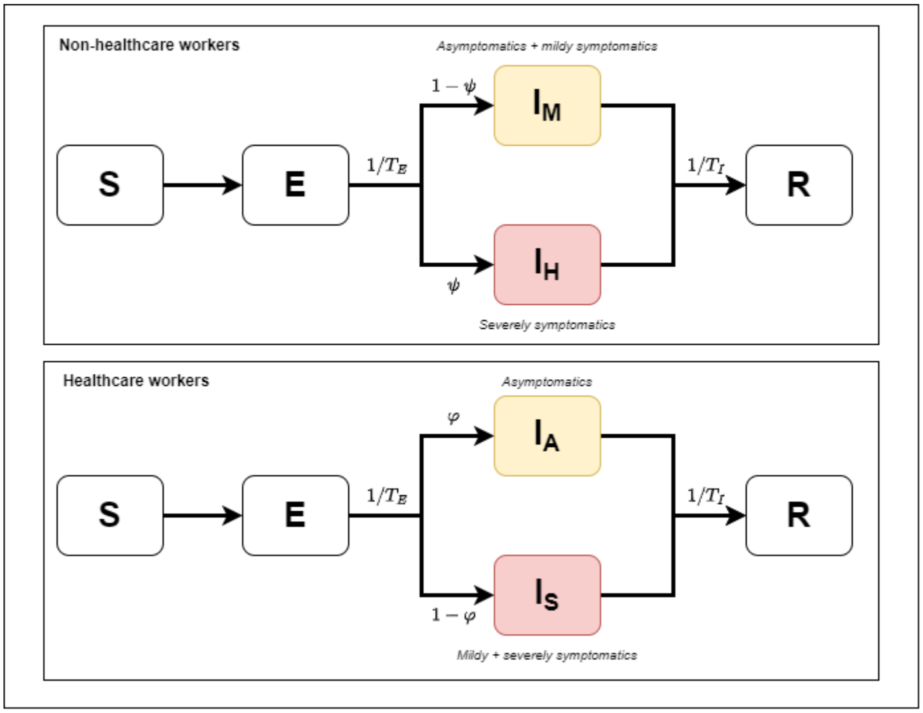


Figure 1: Diagram describing successibe infections statuses for non-HCWs (upper panel) and HCWs (lower panel)

##### In the usual hospital

$$\beta_1^{pp} = \beta_1 \quad (5)$$
$$\beta_1^{ph} = \beta_1 \quad (6)$$
$$\beta_1^{hh} = \frac{\beta_1}{3} \quad (7)$$
$$\beta_1^{h} = \frac{\beta_1}{3} \quad (8)$$
$$(9)$$

Note here that in the reference strategy, $\beta_1=\beta_1^{ref}$ while in the quarantine hospital strategy, $\beta_1=\beta_1^{Q}$.

##### In the quarantine hospital

$$\beta_2^{ph} = \beta_2 \quad (10)$$
$$\beta_2^{hh} = \beta_2 \quad (11)$$

#### Forces of infection

The infectiousness of asymptomatic HCWs is reduced by a factor $\kappa$, as described in the Methods. The average reduction $\kappa^{mild}$ in the infectiousness of non-severe patients (including both asymptomatics and mild symptomatics) may be computed as:

$$\kappa^{mild} = \kappa\frac{\varphi}{1-\psi} + \frac{1-\varphi-\psi}{1-\psi} \quad (12)$$

Forces of infection $FI_X^i$ in setting X for individuals of category i are then computed as follows.

##### In the reference strategy

Infectious individuals include asymptomatic HCWs in the community $(IA_C^h)$ and usual hospital $(IA_1^h)$, symptomatic HCWs isolating in the community $(IS_1^h)$, mildly symptomatic non-HCWs in the community $(IM_C^p)$ and usual hospital $(IM_1^p)$, and severely symptomatic non-HCWs hospitalized in the usual hospital $(IH_1^p)$.
The forces of infection on HCWs may be written as:

$$FI_1^h = \beta_{1_{ref}}^{hh}\kappa\frac{IA_1^h}{N_1^h} + \beta_{1_{ref}}^{ph}\frac{\kappa^{mild}IM_1^p + IH_1^p}{N_1^p} \quad (13)$$
$$FI_C^h = \beta_C^{hh}\left(\kappa\frac{IA_C^h}{N_C^h} + (1-\epsilon)\frac{IS_C^h}{N_C^h}\right) + \beta_C^{ph}\kappa^{mild}\frac{IM_C^p}{N_C^p} \quad (14)$$

The forces of infection on non-HCWs may be written as:

$$FI_1^p = \beta_{1_{ref}}^{hp} \kappa \frac{IA_1^h}{N_1^h} + \beta_{1_{ref}}^{pp} \frac{\kappa^{mild} IM_1^p + IH_1^p}{N_1^p} \tag{15}$$

$$FI_C^p = \beta_C^{hp} \left( \kappa \frac{IA_C^h}{N_C^h} + (1-\epsilon) \frac{IS_C^h}{N_C^h} \right) + \beta_C^{pp} \kappa^{mild} \frac{IM_C^p}{N_C^p} \tag{16}$$

#### In the quarantine hospital strategy

Infectious individuals include asymptomatic HCWs in the community ($IA_C^h$ or $IA_C^L$ if they are self-isolating after a shift in the quarantine hospital), usual hospital ($IA_1^h$) and quarantine hospital, symptomatic HCWs isolating in the community ($IS_1^h$), mildly symptomatic non-HCWs in the community ($IM_C^p$) and usual hospital ($IM_1^p$), and severely symptomatic non-HCWs hospitalized in the quarantine hospital ($IH_2^p$).
The forces of infection on HCWs may be written as:

$$FI_1^h = \beta_1^{hh} \kappa \frac{IA_1^h}{N_1^h} + \beta_1^{ph} \frac{\kappa^{mild} IM_1^p}{N_1^p} \tag{17}$$

$$FI_2^h = \beta_2^{hh} \kappa \frac{IA_2^h}{N_2^h} + \beta_2^{ph} \frac{IH_2^p}{N_2^p} \tag{18}$$

$$FI_C^h = \beta_C^{hh} \times \ contacts + \beta_C^{ph} \kappa^{mild} \frac{IM_C^p}{N_C^p} \tag{19}$$

$$\tag{20}$$

The forces of infection on non-HCWs may be written as:

$$FI_1^p = \beta_1^{hp} \kappa \frac{IA_1^h}{N_1^h} + \beta_1^{pp} \frac{\kappa^{mild} IM_1^p}{N_1^p} \tag{21}$$

$$FI_C^p = \beta_C^{hp} \times \ contacts + \beta_C^{pp} \times \kappa^{mild} \times \frac{IM_C^p}{N_C^p} \tag{22}$$

With infecting contacts with HCWs in the community estimated as:

$$contacts = (1-\epsilon_{H2}) \kappa \frac{IA_C^L}{N_C^h} + \ (1-\epsilon) \frac{IS_C^h}{N_C^h} + \kappa \frac{IA_C^h}{N_C^h} \tag{23}$$

### Hospitalized patient populations

The total hospitalized population remains stable over time, at an assumed "full capacity" computed from the total size of the HCW population $N_{hcw}$ and the patient-to-HCW ratio $R_{ph}$ as:

$$N_{hcw} * R_{ph} * \frac{1}{3} \tag{24}$$

assuming that HCWs spend one third of their time at work in the hospital.

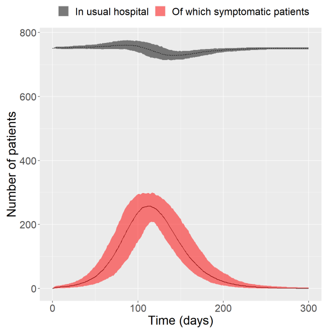


(a) Patient admissions in the usual hospital with the reference strategy

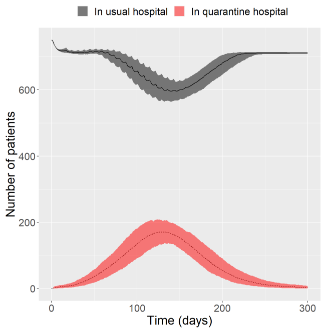


(b) Patient admission in usual and quarantine hospitals with the quarantine hospital strategy

Figure 2: Patient admission in the two scenarios

#### In the reference strategy

In the reference strategy, all patients are hospitalized in the usual hospital, which is assumed to always be filled at capacity. Depending on the stage in the viral epidemic in the community, a more or less large fraction of these patients are hospitalized because they are infected (Fig 2a).

#### In the quarantine hospital strategy

In the quarantine hospital strategy, patients infected with the virus are hospitalized in the quarantine hospital, while others are hospitalized in the usual hospital. As the epidemic progresses and more patients are hospitalized in the quarantine hospital, more HCWs are redirected to work in the quarantine hospitals, and less remain in the usual hospital, leading to less available beds (hence, less hospitalized patients) in the usual hospital (Fig 2b). This reflects for instance de-programming of scheduled surgeries during the COVID-19 pandemic.
